# Cytokine ranking via mutual information algorithm correlates cytokine profiles with presenting disease severity in patients with COVID-19

**DOI:** 10.1101/2020.11.24.20235721

**Authors:** Kelsey E. Huntington, Anna D. Louie, Chun Geun Lee, Jack A. Elias, Eric A. Ross, Wafik S. El-Deiry

## Abstract

Although the range of immune responses to COVID-19 infection is variable, cytokine storm is observed in many affected individuals. To further understand the disease pathogenesis and, consequently, to develop an additional tool for clinicians to evaluate patients for presumptive intervention we sought to compare plasma cytokine levels between a range of donor and patient samples grouped by a COVID-19 Severity Score (CSS) based on need for hospitalization and oxygen requirement. Here we utilize a mutual information algorithm that classifies the information gain for CSS prediction provided by cytokine expression levels and clinical variables. Using this methodology, we found that a small number of clinical and cytokine expression variables are predictive of presenting COVID-19 disease severity, raising questions about the mechanism by which COVID-19 creates severe illness. The variables that were the most predictive of CSS included clinical variables such as age and abnormal chest x-ray as well as cytokines such as macrophage colony-stimulating factor (M-CSF), interferon-inducible protein 10 (IP-10) and Interleukin-1 Receptor Antagonist (IL-1RA). Our results suggest that SARS-CoV-2 infection causes a plethora of changes in cytokine profiles and that particularly in severely ill patients, these changes are consistent with the presence of Macrophage Activation Syndrome and could furthermore be used as a biomarker to predict disease severity.

## INTRODUCTION

In December 2019, severe acute respiratory syndrome coronavirus 2 (SARS-CoV-2), the origin of coronavirus disease 2019 (COVID-19), emerged in Wuhan, China^1^. Although many COVID-19 patients remain asymptomatic, there exists a subset of patients who present with severe illness. Early treatment with dexamethasone appears to improve outcomes in these patients. However, it is not always initially clear which patients would benefit from this therapy^2^. Moreover, COVID-19 infection can be accompanied by a severe inflammatory response characterized by the release of pro-inflammatory cytokines, an event known as cytokine storm (CS) ^3,4^. Thus far, this COVID-19-associated cytokine storm has predominantly been characterized by the presence of IL-1β, IL-2, IL-17, IL-8, TNF, CCL2, and most notably IL-6^3,5–8^. Severe cases of CS can be life-threatening and early diagnosis as well as treatment of this condition can lead to improved outcome. We hypothesize that cytokine profiles combined with clinical information can predict disease severity, potentially giving clinicians an additional tool when evaluating patients for preemptive intervention.

## RESULTS

Analysis was performed for 36 PCR-confirmed COVID-19 (+) and 36 (-) human plasma samples (Source Data Table 1). The COVID-19 Severity Score (CSS) was developed to categorize patients based on their status upon presentation to the emergency department. CSS is graded as follows: 0= COVID(-), No Symptoms, Healthy Control (n=24), 1= COVID(-), Symptoms (n=12), 2= COVID(+), Discharged from Emergency Room (n=15), 3= COVID(+), Admitted, No Oxygen, (n=7), 4= COVID(+), Admitted, Oxygen, (n=8),and 5= COVID(+), Admitted to ICU/Step-Down (n=6) (Figure 1). CSS was used as the outcome variable for a mutual information minimum redundancy maximum relevance algorithm (Figure 1) with the goal of selecting a subset of variables most predictive of CSS. The algorithm confirmed the predictive value of clinical variables such as age and chest x-ray abnormality and also ranked the information gain provided by each of 15 cytokines tested. Several cytokines were able to add unique predictive value to the mutual information model in addition to what was provided by clinical factors such as age or patient comorbidities. This algorithm also deprioritized factors when their predictive value was redundant with the most predictive variables. M-CSF was ranked second after age as it was the factor that added the most predictive power to the algorithm with minimal redundancy with age. It ranked ahead of abnormalities on chest x-ray because while both were relevant in predicting COVID severity, part of the predictiveness of chest x-ray abnormality was also explained by age differences (Figure 3). The top 4 cytokines combined with age were predictive of the most severe CSS (4-5) and had a receiver operating characteristic (Figure 3) with an area under the curve of 0.86. Multiple cytokines, including M-CSF (p<0.01), IP-10 (p<0.01), IL-18 (p=0.01) and IL-1RA (p<0.01) were more relevant in predicting COVID Severity Score than more frequently characterized cytokines in the context of COVID-19 such as IL-6 (p<0.01). These cytokines showed a statistically significant difference in their profiles when segregated by COVID Severity Score (Figure 2), yet the mutual information algorithm prioritized them differently than would be expected based on univariate analyses. This indicates that the mutual information algorithm is prioritizing cytokines whose predictive value for COVID-19 severity cannot be fully explained by other clinical variables such as age or medical comorbidities.

**Figure 1.**
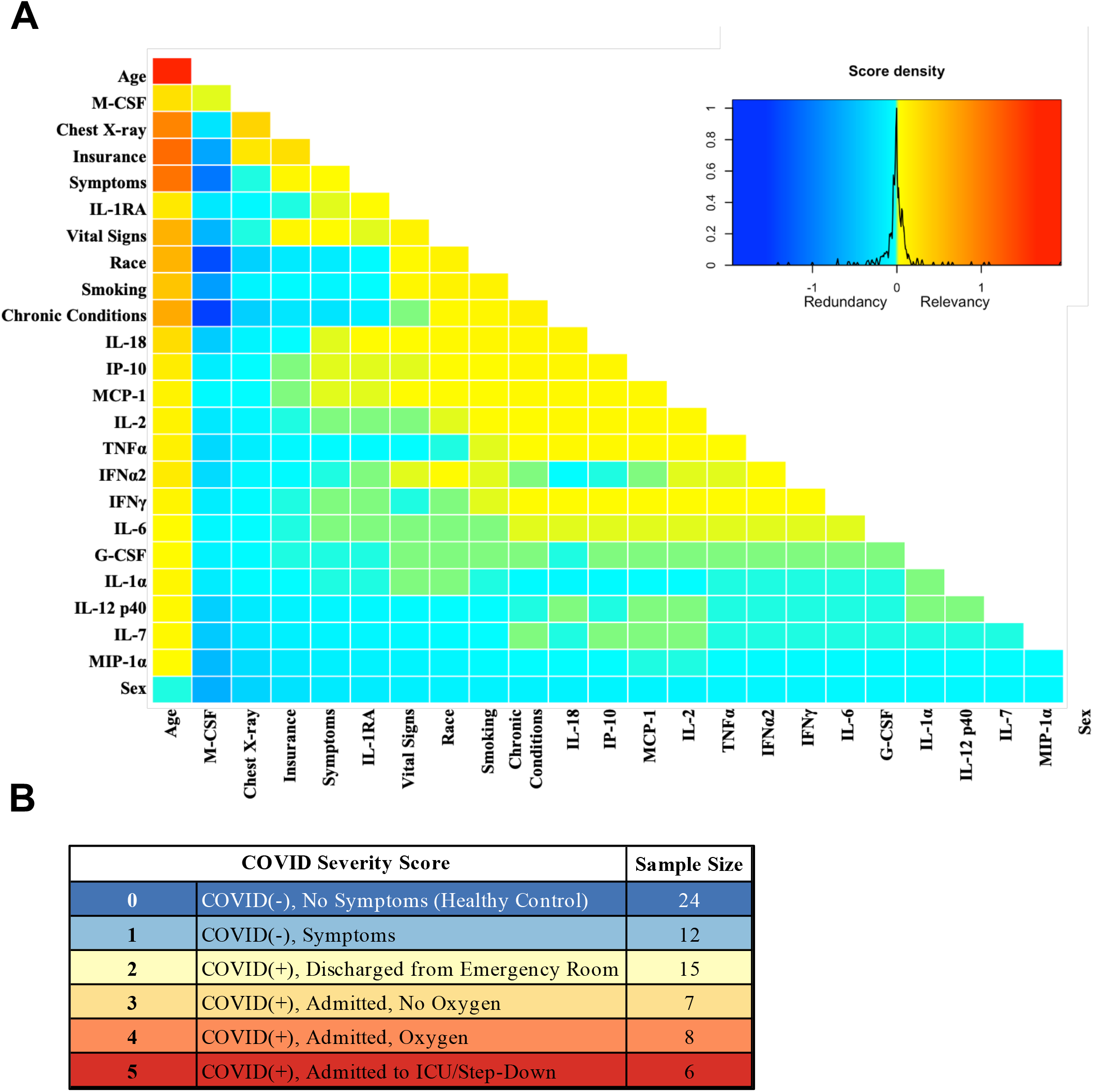
Mutual information COVID Severity Score relevancy matrix. A) Comprehensive matrix of relevancy to CSS of all variables assessed by mutual information algorithm, relevancy scores computed for not-yet selected variables are shown in each column, and variables are ordered to place maximum local scores on the diagonal, yielding a list in decreasing order from the upper left of variable relevancy. Warmer colors indicate higher relevancy while cooler colors indicate higher redundancy. B) COVID Severity Score Table with breakdown of categories as well as sample size per category.

**Figure 2.**
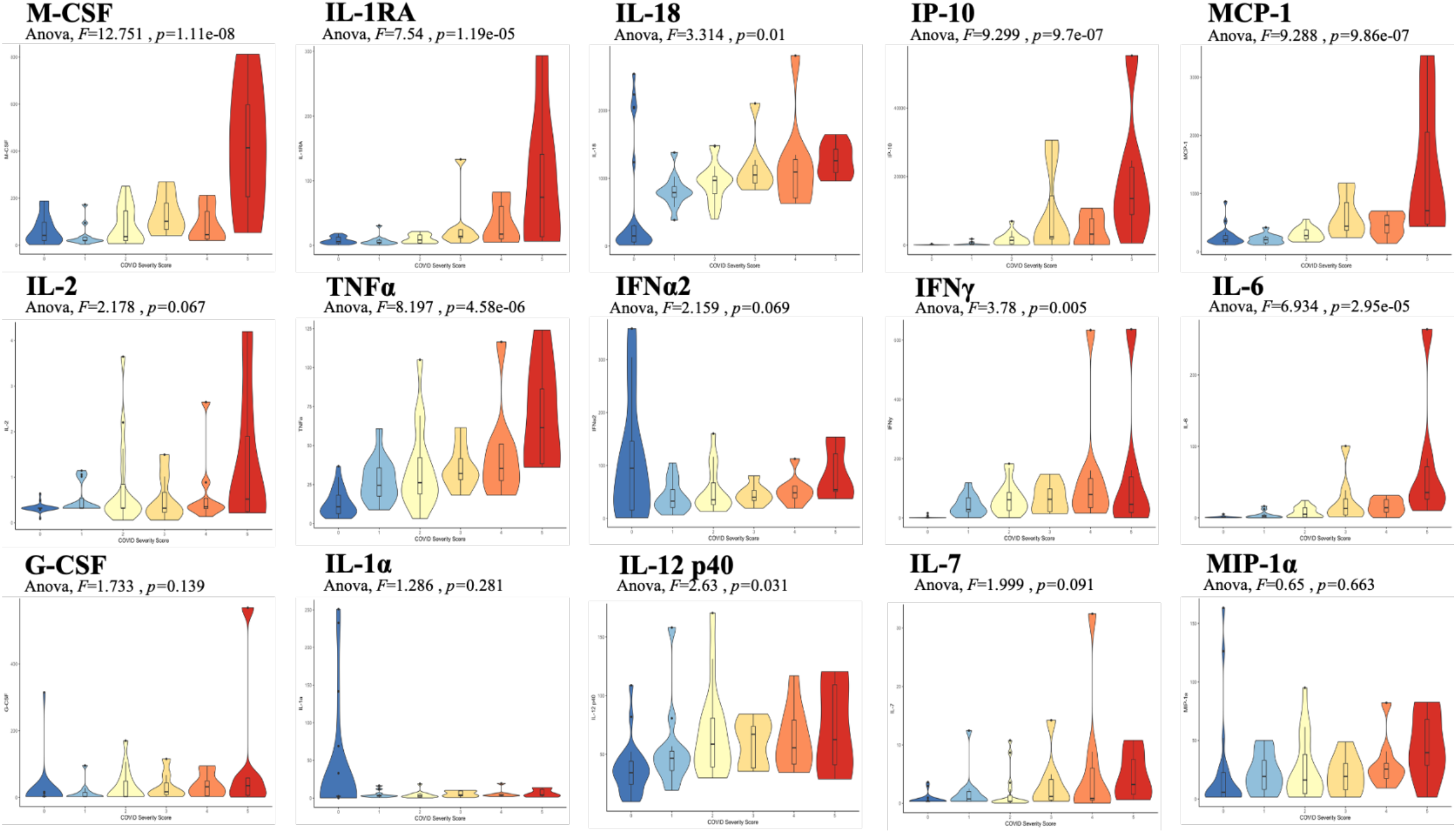
Violin plot representations of cytokine expression levels ordered by COVID Severity Score. Cytokines ordered by row from upper left corner based on mutual information relevancy matrix (upper left being most relevant and lower right being least relevant). The X-axis is CSS and the y-axis is analyte concentration in pg/mL. One-way Anova F values and p values are listed on each plot.

**Figure 3.**
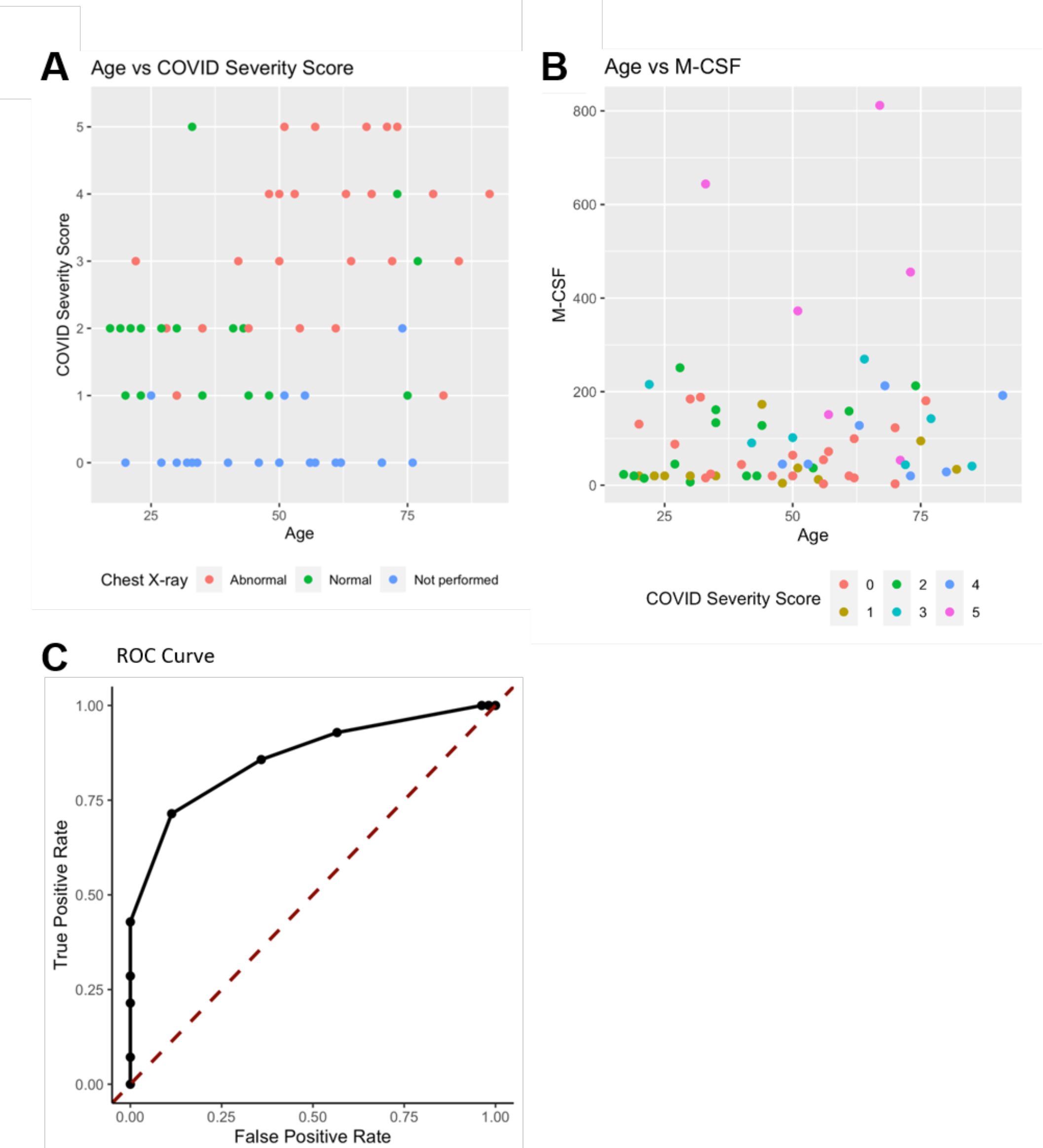
Age, M-CSF, and Chest X-ray are the most predictive variables for COVID Severity Score. A) The y-axis is COVID Severity Score, and the x-axis is age in years with points colored by chest x-ray status. B) The y-axis is age in years and the x-axis is M-CSF concentration in pg/mL, with points colored based on COVID Severity Score. See individual legends below graphs. C) Receiver Operating Characteristic (ROC) Curve predicting CSS 4-5 using Age, M-CSF, IP-10, IL-18, and IL-1RA.

## DISCUSSION

We found that a small number of clinical variables when combined with cytokine expression are predictive of presenting COVID-19 disease severity. Cytokines singled out for relevance by the mutual information algorithm shared a connection to macrophage activation syndrome (MAS), raising questions about the mechanism by which SARS-CoV-2 creates severe illness in a subset of patients. First, we examined the significant contribution of IP-10 to CSS. IP-10 is secreted by monocytes, fibroblasts, and endothelial cells in response to IFN-γ, which is secreted by T cells (mainly, Th1), macrophages, mucosal epithelial cells, and NK cells^9^. This release of IFN-γ induces several cell types to produce IP-10, which consequently recruits more Th1 cells, contributing to a positive feedback loop. IP-10 is also chemoattractant to CXCR3-postitive cells such as macrophages, dendritic cells, NK cells, and T cells. It has been proposed that macrophages recruited by IP-10, in the presence of persistent IFN-γ production, can lead to macrophage activation syndrome (MAS)^5,6,8^. MAS is characterized as a state of systemic hyperinflammation often accompanied by CS which, without intervention, can lead to severe tissue damage and in extreme cases, death^8^.

Moreover, the cytokine most relevant in predicting CSS was M-CSF, which is secreted by eukaryotic cells in response to viral infection and stimulates hematopoietic stem cells to differentiate into macrophages. Currently, there are three separate immune stages that describe the progression of COVID-19. The first stage is characterized by a potent induction of interferons that marks the early activation of the immune system that is important in the viral response and the second stage is characterized by a delayed interferon response^5^. These stages may prime the body for a third stage comprised of detrimental hyperinflammation characterized by CS and MAS^5^. This excessive macrophage activation could explain the increase in IL1-RA that we observed, a cytokine abundantly produced by macrophages.

Steroids have shown a survival benefit for COVID-19, likely by suppressing such detrimental hyperinflammation^2^. Our analysis identified a pattern of cytokine alterations on presentation associated with COVID-19 severity. The ability to identify a cytokine pattern less redundant with known clinical factors such as age and chest x-ray could help better identify patients in need of immunomodulatory treatment without the confounders of current models where the measured cytokines correlate as much with age as with severity^10^. Further studies should be conducted to clarify the mechanistic role that these cytokines and macrophages play in the various stages of COVID-19. The results of these future studies could identify more targeted immunomodulatory strategies beyond steroid administration such as treatment with MEK inhibitors^13^, as well as the ideal timing of these interventions to maximize therapeutic efficacy. Finally, we present the application of this mutual information algorithm as a way to evaluate the dataset as a whole and elucidate the most important cytokines in predicting presenting severity of COVID-19. COVID-19 severity is influenced by many clinical factors, such as age, and this algorithm is able to identify cytokines that contribute information not present in the tested clinical variables. Identifying the most important variables for severe presentation of COVID-19 within a more complete cytokine profile may help determine global immune mechanisms of disease severity.

## METHODS

### Biobank Samples

COVID-19 (+) and (-) human plasma samples were received from the Lifespan Brown COVID-19 Biobank from Brown University at Rhode Island Hospital (Providence, Rhode Island). All patient samples were deidentified but included the available clinical information as described in the manuscript. The IRB study protocol “Pilot Study Evaluating Cytokine Profiles in COVID-19 Patient Samples” did not meet the definition of human subjects research by either the Brown University or the Rhode Island Hospital IRBs. All samples were thawed and centrifuged to remove cellular debris immediately before the assay was run.

### Donor Samples

Normal, healthy, COVID-19 (−) samples were commercially available form Lee BioSolutions (991–58-PS-1, Lee BioSolutions, Maryland Heights, Missouri). All samples were thawed and centrifuged to remove cellular debris immediately before the assay was run.

### Cytokine and chemokine measurements

A MilliPlex MILLIPLEX® MAP Human Cytokine/Chemokine/Growth Factor Panel A-Immunology Multiplex Assay (HCYTA-60K-13, Millipore Sigma, Burlington, Massachusetts) was run on a Luminex 200 Instrument (LX200-XPON-RUO, Luminex Corporation, Austin, Texas) according to the manufacturer’s instructions. Production of granulocyte colony-stimulating factor (G-CSF), interferon gamma (IFNγ), interleukin 1 alpha (IL-1α), interleukin-1 receptor antagonist (IL-1RA), IL-2, IL-6, IL-7, IL-12, interferon-inducible protein 10 (IP-10), monocyte chemoattractant protein-1 (MCP-1), macrophage colony-stimulating factor (M-CSF), macrophage inflammatory protein-1 alpha (MIP-1α), and tumor necrosis factor alpha (TNFα) in the culture supernatant were measured. Data pre-processing: values below limit of detection were re-coded as half the limit of detection. A single extreme outlier value in IFNy levels was removed after confirming outlier status via Hampel and Grubbs outlier testing (both p<0.01).

### Clinical Variables

Available deidentified clinical variables were collected from patients and from chart review during their time in the emergency department. Clinical variables were categorized to create combined variables such as the number of chronic conditions, or the number of presenting symptoms. The full breakdown of clinical variable categorization can be found in Source Data Table 2.

### Data analysis

Data analysis and visualization were generated using R^11^. The varrank package^12^ was used to apply a minimum redundancy maximum relevance mutual information algorithm. The algorithm classifies the amount of information each cytokine and clinical variable can provide about the outcome variable, COVID Severity Score (CSS). Each cytokine variable was discretized into two clusters-either high or low analyte concentration in pg/mL-using k-means clustering to minimize within-variable entropy and, thus, over-fitting. This algorithm partitions each data point into the cluster (high or low analyte concentration) with the nearest mean. Clinical variables and cytokine levels were used to predict CSS. The first variable was selected for local optimum relevance by a greedy algorithm. All subsequent variables were ordered to maximize relevancy and minimize redundancy. The ordering was robust to leave-one-out cross-validation. For each cytokine, one way-ANOVA with Tukey’s honest significant difference test was used to compare plasma cytokine levels among CSS groups.

## Data Availability

All data is contained within the manuscript or supplementary files

## ACKNOWLEDGMENTS

The work was supported by a Brown University COVID-19 Seed Grant (to W.S.E-D.). The COVID-19 Biobank through which plasma samples were obtained was supported by Institutional Development Award Number U54GM115677 from the National Institute of General Medical Sciences of the National Institutes of Health, which funds Advance Clinical and Translational Research (Advance-CTR). The content is solely the responsibility of the authors and does not necessarily represent the official views of the National Institutes of Health. W.S.E-D. is an American Cancer Society Research Professor.

## DISCLOSURES

The authors declare that there are no relevant conflicts of interest.

## Notes

### Competing Interest Statement

The authors have declared no competing interest.

### Author Declarations

COVID-19 (°) and (−) human plasma samples were received from the Lifespan Brown COVID-19 Biobank from Brown University at Rhode Island Hospital (Providence, Rhode Island). All patient samples were deidentified but included the available clinical information as described in the manuscript. The IRB study protocol Pilot Study Evaluating Cytokine Profiles in COVID-19 Patient Samples did not meet the definition of human subjects research by either the Brown University or the Rhode Island Hospital IRBs. This is based on the fact that the project used deidentified specimens from a biobank with a determination that this project did not meet the definition of human subjects research based on specific criteria as described below. The original samples were collected at Rhode Island hospital by the Lifespan Brown COVID-19 Biobank through an IRB-approved protocol that involved informed consent that was used by the biobank. For the present study, we completed a human subjects determination form for the Human Subjects Protection Program at Brown University. We explained the purpose of our research and that we would be receiving deidentified samples from the COVID-19 biobank. We further answered questions about our study that led to the determination that our study constitutes research because we answered "yes" to the following two questions: 1- Does your proposed project involve a systematic investigation: that is a prospective plan that incorporates qualitative or quantitative data collection, and data analysis to answer a question?, and 2- Is the intent of your proposed project to develop or contribute to generalizable knowledge; that is to create knowledge from which conclusions will be drawn that can be applied to populations beyond the specific population from which it was collected. In addition, we answered "no" to four questions regarding whether the project involves human subjects. The questions were 1- Does your proposed project involve an intervention: that is a physical procedure or manipulation or a living individual (or their environment) to obtain information about them? 2- Does your proposed project involve an interaction; that is communication or contact with a living individual (in person, online, or by phone) to obtain information about them? 3-Does your proposed project involve identifiable private information or identifiable biospecimen; that is receipt or collection of private information or biospecimen about a living individual to obtain information about them? and 4- Does your proposed project involve coded information/ biospecimens; that is where a link exists that could allow information about a living individual to be reidentified AND you are able to access the link? Since we answered "no" to all these questions, our proposed project did NOT involve "Human Subjects.” Based on the information included in the Human Subjects Determination Form, The Human Research Protection Program at Brown University agreed with the investigator's self-determination that the project does not meet the definition of human subjects research. This determination was made by the Human Research Protection Program at Brown University on June 17, 2020.

