## Supplementary material for "Cytokine ranking via mutual information algorithm correlates cytokine profiles with presenting disease severity in patients with COVID-19": Source data file

**Source Data Table 1. Patient Information**

| Age  Range | Sex | Race | COVID Severity Score |
| --- | --- | --- | --- |
| 40-49 | Male | Black | 2 |
| 60-69 | Male | Black | 5 |
| 60-69 | Male | Hispanic or Latino | 3 |
| 70-79 | Male | White | 4 |
| 50-59 | Male | Other | 4 |
| 30-39 | Male | White | 2 |
| 50-59 | Male | Other | 5 |
| 60-69 | Male | Asian or Pacific Islander | 2 |
| 10-19 | Female | Unknown | 2 |
| 40-49 | Female | White | 1 |
| 20-29 | Male | Hispanic or Latino | 1 |
| 20-29 | Male | Other | 2 |
| 40-49 | Male | Hispanic or Latino | 2 |
| 50-59 | Male | Other | 1 |
| 30-39 | Male | Black | 1 |
| 30-39 | Female | Other | 2 |
| 40-49 | Male | Hispanic or Latino | 4 |
| 50-59 | Male | Hispanic or Latino | 1 |
| 30-39 | Female | White | 2 |
| 30-39 | Male | Hispanic or Latino | 5 |
| 70-79 | Female | White | 5 |
| 50-59 | Female | Asian or Pacific Islander | 2 |
| 30-39 | Male | Other | 1 |
| 70-79 | Female | Hispanic or Latino | 2 |
| 50-59 | Female | Hispanic or Latino | 4 |
| 80-89 | Female | Hispanic or Latino | 3 |
| 80-89 | Male | Hispanic or Latino | 4 |
| 70-79 | Male | Hispanic or Latino | 3 |
| 40-49 | Female | Hispanic or Latino | 3 |
| 80-89 | Male | Black | 1 |
| 20-29 | Male | Hispanic or Latino | 2 |
| 20-29 | Female | Black | 1 |
| 20-29 | Male | Hispanic or Latino | 1 |
| 20-29 | Male | Hispanic or Latino | 2 |
| 70-79 | Male | Hispanic or Latino | 3 |
| 70-79 | Male | White | 1 |
| 90-99 | Female | White | 4 |
| 40-49 | Female | Hispanic or Latino | 2 |
| 50-59 | Male | Hispanic or Latino | 3 |
| 10-19 | Male | Hispanic or Latino | 2 |
| 60-69 | Female | Hispanic or Latino | 4 |
| 70-79 | Male | White | 5 |
| 30-39 | Female | White | 1 |
| 20-29 | Female | Hispanic or Latino | 3 |
| 50-59 | Male | Hispanic or Latino | 5 |
| 20-29 | Female | Asian or Pacific Islander | 2 |
| 40-49 | Female | Black | 1 |
| 60-69 | Female | White | 4 |
| 50-59 | Female | White | 0 |
| 40-49 | Female | White | 0 |
| 30-39 | Male | White | 0 |
| 60-69 | Female | White | 0 |
| 50-59 | Male | Asian | 0 |
| 70-79 | Male | White | 0 |
| 20-29 | Female | White | 0 |
| 30-39 | Male | Asian | 0 |
| 70-79 | Female | White | 0 |
| 20-29 | Female | White | 0 |
| 50-59 | Female | White | 0 |
| 50-59 | Male | White | 0 |
| 60-69 | Male | White | 0 |
| 70-79 | Male | White | 0 |
| 30-39 | Female | White | 0 |
| 60-69 | Male | Hispanic | 0 |
| 30-39 | Female | Hispanic | 0 |
| 40-49 | Male | White | 0 |
| 50-59 | Female | White | 0 |

**Source Data Table 2. Mutual Algorithm Criteria Table**

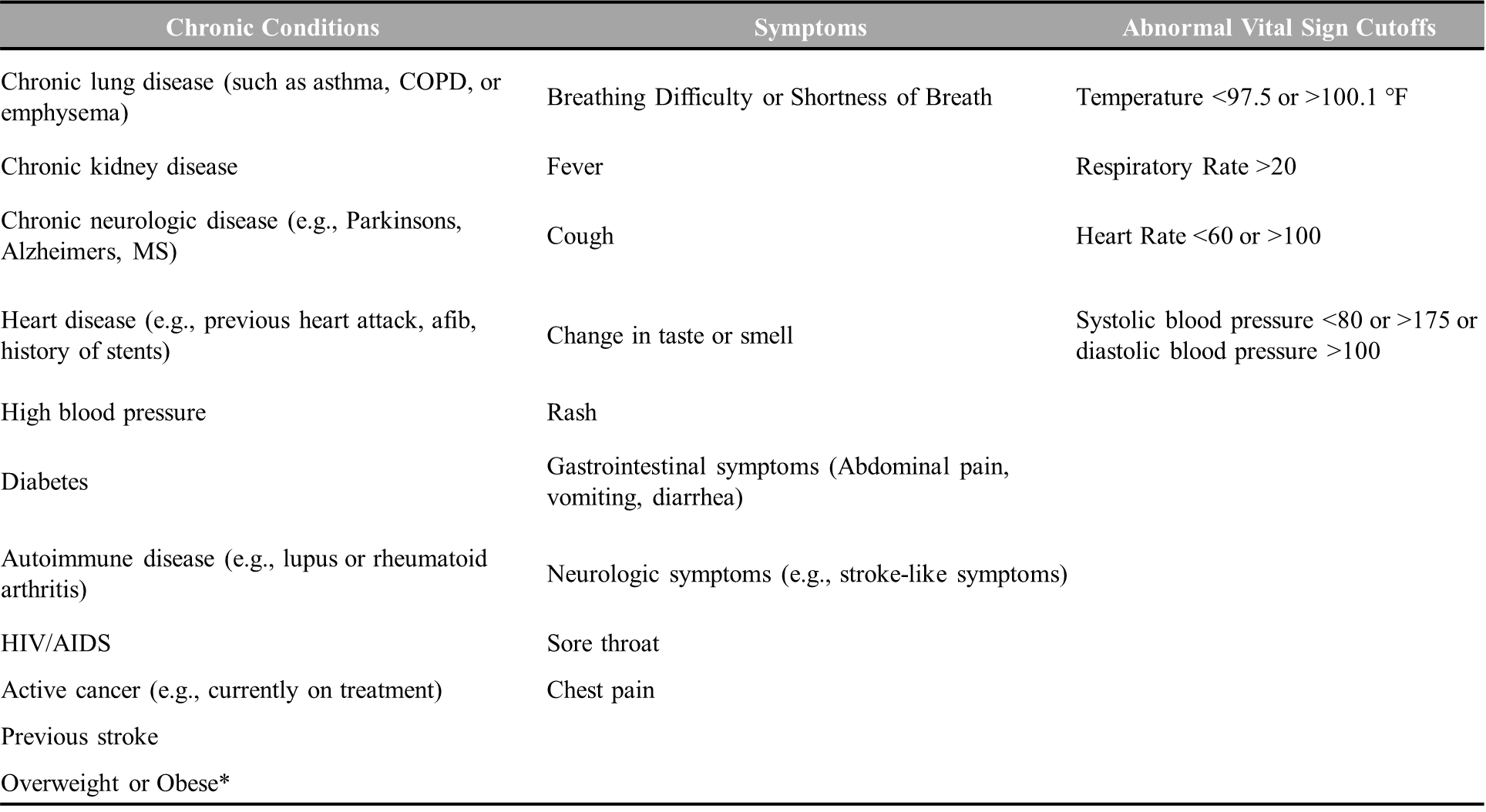

*documented obesity or overweight for height <99th percentile
